## Supplementary Material for "Burden of Post-COVID-19 Syndrome and Implications for Healthcare Service Planning: A Population-based Cohort Study"

---

**Supplementary Material**

---

**Table S1**: Results from univariable and multivariable logistic regression models for the outcome of not having fully recovered at six to eight months after diagnosis.

| Variable | | Univariable | | | Multivariable [a] | | |
|---|---|---|---|---|---|---|---|
| | N | OR | 95% CI | p-value | OR | 95% CI | p-value |
| **Age group (years)** | *385* | | | | | | |
| 18-39 | | — | — | | — | — | |
| 40-64 | | 1.96 | 1.20 to 3.25 | 0.008 | 1.59 | 0.93 to 2.73 | 0.093 |
| ≥65 | | 1.45 | 0.70 to 2.95 | 0.31 | 0.97 | 0.41 to 2.20 | 0.94 |
| **Sex** | *385* | | | | | | |
| Female | | — | — | | — | — | |
| Male | | 0.59 | 0.38 to 0.92 | 0.02 | 0.53 | 0.33 to 0.85 | 0.009 |
| **Time since diagnosis (days)** | *385* | 1.00 | 0.99 to 1.00 | 0.69 | 0.99 | 0.99 to 1.00 | 0.10 |
| **Initial symptom severity** | *385* | | | | | | |
| Mild to moderate | | — | — | | — | — | |
| Severe to very severe | | 2.5 | 1.60 to 3.94 | <0.001 | 2.05 | 1.27 to 3.34 | 0.003 |
| **Initial hospitalization** | *385* | | | | | | |
| No | | — | — | | — | — | |
| Yes | | 1.75 | 1.03 to 2.96 | 0.038 | 1.17 | 0.63 to 2.16 | 0.61 |
| **Initial ICU stay** | *385* | | | | | | |
| No | | — | — | | — | — | |
| Yes | | 1.06 | 0.23 to 3.89 | 0.93 | 0.55 | 0.10 to 2.49 | 0.45 |
| **Smoking status** | *382* | | | | | | |
| Non-smoker | | — | — | | — | — | |
| Ex-smoker | | 1.44 | 0.88 to 2.36 | 0.15 | 1.48 | 0.87 to 2.52 | 0.14 |
| Smoker | | 1.21 | 0.61 to 2.32 | 0.56 | 1.61 | 0.78 to 3.24 | 0.19 |
| **Body mass index** | *379* | 1.07 | 1.02 to 1.12 | 0.004 | 1.04 | 0.99 to 1.09 | 0.15 |
| **Comorbidities** | *384* | | | | | | |
| No | | — | — | | — | — | |
| Yes | | 2.21 | 1.40 to 3.48 | <0.001 | 2.08 | 1.24 to 3.50 | 0.005 |

*Legend: OR = Odds Ratio, CI = Confidence Interval, ICU = Intensive Care Unit; [a] adjusted for age group, sex, initial hospitalization, symptom severity, and comorbidities.*

**Table S2**: Results from univariable and multivariable logistic regression models for the outcome of fatigue at six to eight months after diagnosis.

| Variable | | Univariable | | | Multivariable [a] | | |
|---|---|---|---|---|---|---|---|
| | N | OR | 95% CI | p-value | OR | 95% CI | p-value |
| **Age group (years)** | *426* | | | | | | |
| 18-39 | | — | — | | — | — | |
| 40-64 | | 0.58 | 0.38 to 0.89 | 0.012 | 0.59 | 0.39 to 0.91 | 0.018 |
| ≥65 | | 0.4 | 0.21 to 0.73 | 0.003 | 0.41 | 0.21 to 0.78 | 0.007 |
| **Sex** | *426* | | | | | | |
| Female | | — | — | | — | — | |
| Male | | 0.69 | 0.47 to 1.02 | 0.062 | 0.72 | 0.49 to 1.07 | 0.10 |
| **Time since diagnosis (days)** | *426* | 1.00 | 1.00 to 1.01 | 0.85 | 1.00 | 1.00 to 1.01 | 0.52 |
| **Initial symptom severity** | *426* | | | | | | |
| Asymptomatic | | — | — | | — | — | |
| Mild to moderate | | 0.99 | 0.52 to 1.86 | 0.96 | 0.96 | 0.50 to 1.83 | 0.89 |
| Severe to very severe | | 1.34 | 0.69 to 2.59 | 0.38 | 1.38 | 0.69 to 2.73 | 0.36 |
| **Initial hospitalization** | *426* | | | | | | |
| No | | — | — | | — | — | |
| Yes | | 0.77 | 0.47 to 1.26 | 0.30 | 1.00 | 0.59 to 1.71 | 0.99 |
| **Initial ICU stay** | *426* | | | | | | |
| No | | — | — | | — | — | |
| Yes | | 2.96 | 0.71 to 20.0 | 0.18 | 4.63 | 1.02 to 32.9 | 0.07 |
| **Smoking status** | *424* | | | | | | |
| Non-smoker | | — | — | | — | — | |
| Ex-smoker | | 1.35 | 0.87 to 2.11 | 0.18 | 1.58 | 1.00 to 2.52 | 0.05 |
| Smoker | | 1.47 | 0.83 to 2.63 | 0.19 | 1.27 | 0.71 to 2.31 | 0.42 |
| **Body mass index** | *419* | 1.01 | 0.97 to 1.05 | 0.52 | 1.04 | 1.00 to 1.09 | 0.08 |
| **Comorbidities** | *426* | | | | | | |
| No | | — | — | | — | — | |
| Yes | | 0.91 | 0.61 to 1.36 | 0.65 | 1.27 | 0.81 to 2.01 | 0.30 |

*Legend: OR = Odds Ratio, CI = Confidence Interval, ICU = Intensive Care Unit; [a] adjusted for age group, sex, and initial hospitalization.*

**Table S3**: Results from univariable and multivariable logistic regression models for the outcome of mMRC dyspnea grade ≥1 at six to eight months after diagnosis.

| Variable | | Univariable | | | Multivariable [a] | | |
|---|---|---|---|---|---|---|---|
| | N | OR | 95% CI | p-value | OR | 95% CI | p-value |
| **Age group (years)** | *395* | | | | | | |
| 18-39 | | — | — | | — | — | |
| 40-64 | | 1.61 | 0.96 to 2.76 | 0.077 | 0.79 | 0.41 to 1.49 | 0.46 |
| ≥65 | | 2.88 | 1.45 to 5.73 | 0.002 | 0.90 | 0.37 to 2.16 | 0.82 |
| **Sex** | *395* | | | | | | |
| Female | | — | — | | — | — | |
| Male | | 0.59 | 0.37 to 0.94 | 0.029 | 0.45 | 0.26 to 0.76 | 0.003 |
| **Time since diagnosis (days)** | *395* | 1.00 | 0.99 to 1.01 | 0.74 | 1.00 | 0.99 to 1.01 | 0.92 |
| **Initial symptom severity** | *395* | | | | | | |
| Asymptomatic | | — | — | | — | — | |
| Mild to moderate | | 0.87 | 0.40 to 2.07 | 0.74 | 1.00 | 0.40 to 2.73 | >0.99 |
| Severe to very severe | | 1.93 | 0.89 to 4.54 | 0.11 | 1.42 | 0.57 to 3.87 | 0.47 |
| **Initial hospitalization** | *395* | | | | | | |
| No | | — | — | | — | — | |
| Yes | | 4.06 | 2.39 to 6.91 | <0.001 | 4.17 | 2.23 to 7.91 | <0.001 |
| **Initial ICU stay** | *395* | | | | | | |
| No | | — | — | | — | — | |
| Yes | | 4.05 | 1.05 to 16.7 | 0.04 | 1.05 | 0.22 to 5.31 | 0.95 |
| **Smoking status** | *393* | | | | | | |
| Non-smoker | | — | — | | — | — | |
| Ex-smoker | | 1.62 | 0.97 to 2.69 | 0.06 | 1.66 | 0.92 to 3.00 | 0.093 |
| Smoker | | 0.88 | 0.41 to 1.78 | 0.73 | 1.31 | 0.57 to 2.85 | 0.51 |
| **Body mass index** | *388* | 1.13 | 1.08 to 1.20 | <0.001 | 1.14 | 1.08 to 1.20 | <0.001 |
| **Respiratory condition** | *388* | | | | | | |
| No | | — | — | | — | — | |
| Yes | | 2.36 | 1.10 to 4.95 | 0.024 | 1.71 | 0.70 to 4.01 | 0.22 |
| **Comorbidities** | *395* | | | | | | |
| No | | — | — | | — | — | |
| Yes | | 3.93 | 2.44 to 6.40 | <0.001 | 2.71 | 1.38 to 5.36 | 0.004 |

*Legend: OR = Odds Ratio, CI = Confidence Interval, ICU = Intensive Care Unit; [a] adjusted for age group, sex, initial hospitalization, smoking, respiratory comorbidity, and body mass index.*

**Table S4:** Results from univariable and multivariable logistic regression models for the outcome of depression at six to eight months after diagnosis.

| Variable | | Univariable | | | Multivariable [a] | | |
|---|---|---|---|---|---|---|---|
| | N | OR | 95% CI | p-value | OR | 95% CI | p-value |
| **Age group (years)** | *428* | | | | | | |
| 18-39 | | — | — | | — | — | |
| 40-64 | | 1.05 | 0.66 to 1.69 | 0.83 | 0.97 | 0.59 to 1.59 | 0.91 |
| ≥65 | | 1.19 | 0.60 to 2.28 | 0.61 | 1.18 | 0.56 to 2.42 | 0.65 |
| **Sex** | *428* | | | | | | |
| Female | | — | — | | — | — | |
| Male | | 0.70 | 0.45 to 1.08 | 0.11 | 0.74 | 0.47 to 1.15 | 0.18 |
| **Time since diagnosis (days)** | *428* | 1.00 | 0.99 to 1.00 | 0.65 | 1.00 | 0.99 to 1.00 | 0.28 |
| **Initial symptom severity** | *428* | | | | | | |
| Asymptomatic | | — | — | | — | — | |
| Mild to moderate | | 0.84 | 0.40 to 1.92 | 0.67 | 0.85 | 0.40 to 1.94 | 0.69 |
| Severe to very severe | | 2.08 | 0.99 to 4.71 | 0.062 | 2.05 | 0.96 to 4.69 | 0.074 |
| **Initial hospitalization** | *428* | | | | | | |
| No | | — | — | | — | — | |
| Yes | | 1.42 | 0.82 to 2.39 | 0.20 | 1.05 | 0.57 to 1.89 | 0.88 |
| **Initial ICU stay** | *428* | | | | | | |
| No | | — | — | | — | — | |
| Yes | | 0.81 | 0.12 to 3.42 | 0.80 | 0.47 | 0.07 to 2.17 | 0.37 |
| **Smoking status** | *426* | | | | | | |
| Non-smoker | | — | — | | — | — | |
| Ex-smoker | | 1.18 | 0.72 to 1.94 | 0.51 | 1.21 | 0.72 to 2.01 | 0.48 |
| Smoker | | 1.56 | 0.83 to 2.84 | 0.16 | 1.71 | 0.89 to 3.21 | 0.10 |
| **Body mass index** | *421* | 1.02 | 0.98 to 1.06 | 0.40 | 1.02 | 0.97 to 1.06 | 0.51 |
| **Comorbidities** | *428* | | | | | | |
| No | | — | — | | — | — | |
| Yes | | 1.48 | 0.94 to 2.31 | 0.086 | 1.41 | 0.84 to 2.34 | 0.19 |
| **Education** | *426* | | | | | | |
| None or mandatory school | | — | — | | — | — | |
| Vocational training or specialized baccalaureate | | 0.34 | 0.13 to 0.90 | 0.028 | 0.31 | 0.11 to 0.85 | 0.022 |
| Higher technical school or college | | 0.28 | 0.10 to 0.76 | 0.013 | 0.27 | 0.09 to 0.79 | 0.016 |
| University | | 0.25 | 0.09 to 0.68 | 0.006 | 0.23 | 0.08 to 0.65 | 0.006 |
| **Employment** | *424* | | | | | | |
| Employed | | — | — | | — | — | |
| Student | | 0.53 | 0.08 to 2.01 | 0.42 | 0.6 | 0.09 to 2.41 | 0.53 |

| | | | | | | | |
|---|---|---|---|---|---|---|---|
| Retired | | 1.41 | 0.76 to 2.54 | 0.26 | 1.89 | 0.58 to 6.11 | 0.28 |
| Unemployed or other | | 2.60 | 1.18 to 5.64 | 0.016 | 2.53 | 1.12 to 5.63 | 0.023 |
| **Income** | *405* | | | | | | |
| <6'000 CHF | | — | — | | — | — | |
| 6'000 - 12'000 CHF | | 0.91 | 0.54 to 1.53 | 0.72 | 1.05 | 0.61 to 1.81 | 0.87 |
| >12'000 CHF | | 0.71 | 0.39 to 1.25 | 0.24 | 0.84 | 0.45 to 1.55 | 0.57 |

*Legend: OR = Odds Ratio, CI = Confidence Interval, ICU = Intensive Care Unit; [a] adjusted for age group, sex, initial hospitalization, and initial symptom severity.*

**Figure S1**: Venn diagram of the overlap of participants that have not fully recovered or are experiencing fatigue, dyspnea or depression at six to eight months after SARS-CoV-2 infection (total N=296).

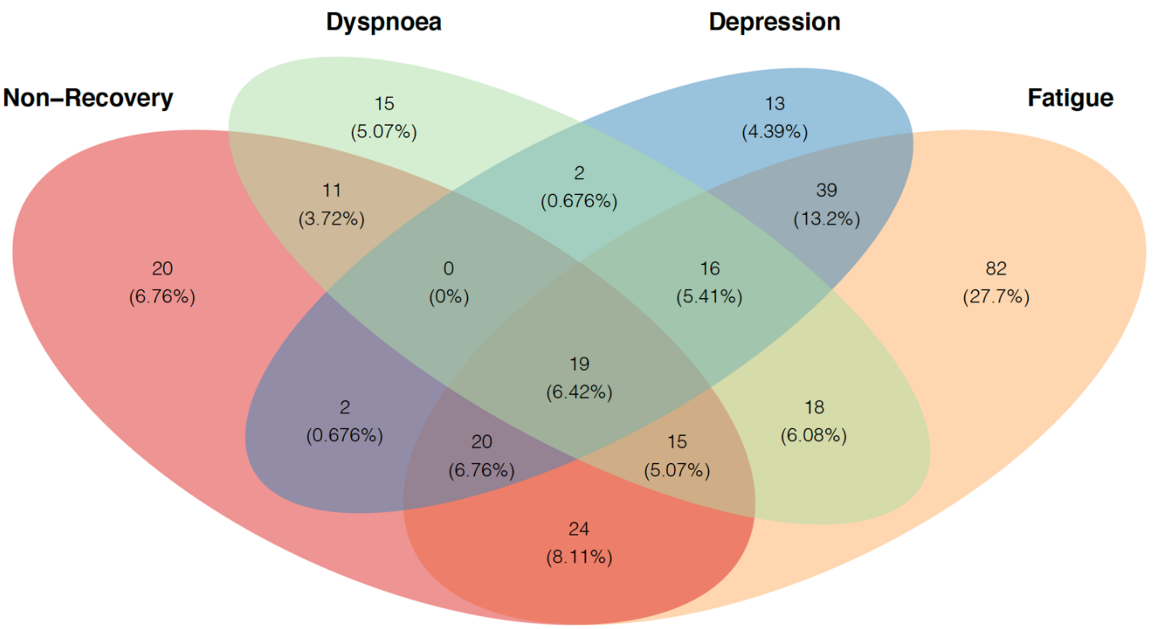

**Table S5**: Overlap of participants not having recovered or experiencing fatigue, dyspnea, or depression and healthcare use at six to eight months after diagnosis.

| | **Healthcare use related to COVID-19** [a,b] | | |
|---|---|---|---|
| **Variable** | **No**, N=251 | **Yes**, N=170 | **Overall**, N=431 |
| **Recovery** | | | |
| Recovered to normal health status | 214 (68.8%) | 97 (31.2%) | 320 (74.2%) |
| Not recovered to normal health status | 37 (33.6%) | 73 (66.4%) | 111 (25.8%) |
| **Fatigue (measured by FAS)** | | | |
| No | 121 (64.0%) | 68 (36.0%) | 193 (45.3%) |
| Yes | 128 (56.1%) | 100 (43.9%) | 233 (54.7%) |
| *Missing* | *2* | *2* | *5* |
| **Dyspnea (measured by mMRC scale)** | | | |
| mMRC grade 0 | 196 (67.4%) | 95 (32.6%) | 299 (75.7%) |
| mMRC grade ≥1 | 36 (37.9%) | 59 (62.1%) | 96 (24.3%) |
| *Missing* | *19* | *16* | *36* |
| **Depression (measured by DASS-21)** | | | |
| No | 203 (65.3%) | 108 (34.7%) | 317 (74.1%) |
| Yes | 48 (44.4%) | 60 (55.6%) | 111 (25.9%) |
| *Missing* | *0* | *2* | *3* |

*Legend: FAS = Fatigue Assessment Scale, mMRC = modified Medical Research Council, DASS-21 = Depression, Anxiety and Stress Score (21 items).* [a] *Data on healthcare use missing from 10 individuals,* [b] *row percentages presented.*

**Table S6**: Results from univariable and multivariable logistic regression models for the outcome of having at least one further healthcare contact (defined as rehospitalization, general practitioner visit or medical hotline call) related to COVID-19 within six to eight months after diagnosis.

| Variable | | Univariable | | | Multivariable [a] | | |
|---|---|---|---|---|---|---|---|
| | N | OR | 95% CI | p-value | OR | 95% CI | p-value |
| **Age group (years)** | *421* | | | | | | |
| 18-39 | | — | — | | — | — | |
| 40-64 | | 2.24 | 1.44 to 3.50 | <0.001 | 1.94 | 1.22 to 3.10 | 0.005 |
| ≥65 | | 3.18 | 1.73 to 5.92 | <0.001 | 2.23 | 1.12 to 4.45 | 0.022 |
| **Sex** | *421* | | | | | | |
| Female | | — | — | | — | — | |
| Male | | 0.67 | 0.45 to 0.99 | 0.047 | 0.62 | 0.41 to 0.95 | 0.03 |
| **Time since diagnosis (days)** | *421* | 1.00 | 1.00 to 1.01 | 0.74 | 1.00 | 0.99 to 1.00 | 0.26 |
| **Initial symptom severity** | *421* | | | | | | |
| Asymptomatic | | — | — | | — | — | |
| Mild to moderate | | 1.46 | 0.72 to 3.19 | 0.31 | 1.50 | 0.72 to 3.34 | 0.30 |
| Severe to very severe | | 3.38 | 1.64 to 7.45 | 0.002 | 2.59 | 1.21 to 5.87 | 0.018 |
| **Initial hospitalization** | *421* | | | | | | |
| No | | — | — | | — | — | |
| Yes | | 4.21 | 2.52 to 7.21 | <0.001 | 2.94 | 1.66 to 5.32 | <0.001 |
| **Initial ICU stay** | *421* | | | | | | |
| No | | — | — | | — | — | |
| Yes | | 14 | 2.59 to 259 | 0.013 | 3.65 | 0.61 to 70.1 | 0.24 |
| **Recovery** | *421* | | | | | | |
| Recovered to normal health status | | — | — | | — | — | |
| Not recovered to normal health status | | 4 | 2.76 to 6.97 | <0.001 | 3.53 | 2.14 to 5.86 | <0.001 |
| **Fatigue** | *417* | | | | | | |
| No | | — | — | | — | — | |
| Yes | | 1 | 0.94 to 2.07 | 0.1 | 1.61 | 1.04 to 2.50 | 0.032 |
| **Dyspnea** | *417* | | | | | | |
| mMRC grade 0 | | — | — | | — | — | |
| mMRC grade ≥1 | | 1 | 0.94 to 2.07 | 0.1 | 2.35 | 1.40 to 3.99 | 0.001 |
| **Depression** | *419* | | | | | | |
| No | | — | — | | — | — | |
| Yes | | 2 | 1.51 to 3.68 | <0.001 | 2.13 | 1.32 to 3.45 | 0.002 |
| **Smoking status** | *418* | | | | | | |
| Non-smoker | | — | — | | — | — | |

| | | | | | | | |
|---|---|---|---|---|---|---|---|
| Ex-smoker | | 1.23 | 0.79 to 1.91 | 0.36 | 1.14 | 0.70 to 1.84 | 0.59 |
| Smoker | | 0.86 | 0.47 to 1.54 | 0.61 | 1.16 | 0.61 to 2.18 | 0.64 |
| **Body mass index** | *413* | 1.01 | 0.98 to 1.05 | 0.48 | 0.99 | 0.94 to 1.03 | 0.57 |
| **Comorbidities** | *420* | | | | | | |
| No | | — | — | | — | — | |
| Yes | | 1.80 | 1.20 to 2.72 | 0.005 | 1.14 | 0.70 to 1.84 | 0.59 |

*Legend: OR = Odds Ratio, CI = Confidence Interval, ICU = Intensive Care Unit; [a] adjusted for age group, sex, initial hospitalization, and initial symptom severity.*

**Table S7**: Sensitivity analysis of relative health status, fatigue, dyspnea, mental health, and health-related quality of life in study participants at six to eight months after SARS-CoV-2 infection, stratified into time periods of limited and increased testing for SARS-CoV-2, as well as with limited and increased awareness of post-COVID-19 syndrome.

| | Test timing (25 Jun 2020) | | Awareness (09 Nov 2020) | | |
|---|---|---|---|---|---|
| **Variable** | **Limited testing**, N=313 | **Increased testing**, N=117 | **Lower awareness of post-COVID syndrome**, N=129 | **Increased awareness of post-COVID syndrome**, N=302 | **Overall**, N=431 |
| **Recovery** | | | | | |
| Recovered to normal health status | 237 (75.7%) | 82 (70.1%) | 102 (79.1%) | 218 (72.2%) | 320 (74.2%) |
| Not recovered to normal health status | 76 (24.3%) | 35 (29.9%) | 27 (20.9%) | 84 (27.8%) | 111 (25.8%) |
| **Self-reported symptoms** | | | | | |
| No new or ongoing symptoms | 235 (75.1%) | 89 (76.1%) | 95 (73.6%) | 230 (76.2%) | 325 (75.4%) |
| Any new or ongoing symptoms | 78 (24.9%) | 28 (23.9%) | 34 (26.4%) | 72 (23.8%) | 106 (24.6%) |
| **Recovery and symptoms** | | | | | |
| Recovered and symptom-free | 197 (62.9%) | 67 (57.3%) | 83 (64.3%) | 182 (60.3%) | 265 (61.5%) |
| Not recovered or experiencing symptoms | 116 (37.1%) | 50 (42.7%) | 46 (35.7%) | 120 (39.7%) | 166 (38.5%) |
| **Fatigue (measured by FAS)** | | | | | |
| No fatigue | 144 (46.8%) | 49 (41.9%) | 69 (54.3%) | 124 (41.5%) | 193 (45.3%) |
| Fatigue | 164 (53.2%) | 68 (58.1%) | 58 (45.7%) | 175 (58.5%) | 233 (54.7%) |
| *Missing* | *5* | *0* | *2* | *3* | *5* |
| **Dyspnea (measured by mMRC scale)** | | | | | |
| mMRC grade 0 | 212 (74.4%) | 86 (78.9%) | 87 (73.1%) | 212 (76.8%) | 299 (75.7%) |
| mMRC grade 1 | 59 (20.7%) | 22 (20.2%) | 29 (24.4%) | 52 (18.8%) | 81 (20.5%) |
| mMRC grade ≥2 | 14 (4.9%) | 1 (0.9%) | 3 (2.5%) | 12 (4.3%) | 15 (3.8%) |
| *Missing* | *28* | *8* | *10* | *26* | *36* |
| **Depression (measured by DASS-21)** | | | | | |
| No depression | 233 (75.2%) | 83 (70.9%) | 99 (77.3%) | 218 (72.7%) | 317 (74.1%) |
| Mild to moderate depression | 59 (19.0%) | 26 (22.2%) | 18 (14.1%) | 67 (22.3%) | 85 (19.9%) |
| Severe to very severe depression | 18 (5.8%) | 8 (6.8%) | 11 (8.6%) | 15 (5.0%) | 26 (6.1%) |
| *Missing* | *3* | *0* | *1* | *2* | *3* |
| **Anxiety (measured by DASS-21)** | | | | | |
| No anxiety | 213 (69.2%) | 77 (65.8%) | 93 (73.2%) | 198 (66.2%) | 291 (68.3%) |
| Mild to moderate anxiety | 68 (22.1%) | 35 (29.9%) | 26 (20.5%) | 77 (25.8%) | 103 (24.2%) |
| Severe to very severe anxiety | 27 (8.8%) | 5 (4.3%) | 8 (6.3%) | 24 (8.0%) | 32 (7.5%) |
| *Missing* | *5* | *0* | *2* | *3* | *5* |
| **Stress (measured by DASS-21)** | | | | | |

| | | | | | |
|---|---|---|---|---|---|
| No stress | 263 (85.7%) | 93 (79.5%) | 112 (88.9%) | 245 (81.9%) | 357 (84.0%) |
| Mild to moderate stress | 33 (10.7%) | 18 (15.4%) | 11 (8.7%) | 40 (13.4%) | 51 (12.0%) |
| Severe to very severe stress | 11 (3.6%) | 6 (5.1%) | 3 (2.4%) | 14 (4.7%) | 17 (4.0%) |
| *Missing* | *6* | *0* | *3* | *3* | *6* |
| **EQ-5D mobility** | | | | | |
| No mobility problems | 276 (88.7%) | 104 (88.9%) | 115 (89.1%) | 266 (88.7%) | 381 (88.8%) |
| Mobility problems | 35 (11.3%) | 13 (11.1%) | 14 (10.9%) | 34 (11.3%) | 48 (11.2%) |
| *Missing* | *2* | *0* | *0* | *2* | *2* |
| **EQ-5D self care** | | | | | |
| No problems with self-care | 310 (99.4%) | 117 (100.0%) | 129 (100.0%) | 299 (99.3%) | 428 (99.5%) |
| Problems with self-care | 2 (0.6%) | 0 (0.0%) | 0 (0.0%) | 2 (0.7%) | 2 (0.5%) |
| *Missing* | *1* | *0* | *0* | *1* | *1* |
| **EQ-5D usual activities** | | | | | |
| No problems during usual activities | 278 (89.1%) | 106 (90.6%) | 117 (90.7%) | 268 (89.0%) | 385 (89.5%) |
| Problems during usual activities | 34 (10.9%) | 11 (9.4%) | 12 (9.3%) | 33 (11.0%) | 45 (10.5%) |
| *Missing* | *1* | *0* | *0* | *1* | *1* |
| **EQ-5D pain & discomfort** | | | | | |
| No pain or discomfort present | 192 (61.7%) | 84 (72.4%) | 84 (65.1%) | 193 (64.5%) | 277 (64.7%) |
| Pain or discomfort present | 119 (38.3%) | 32 (27.6%) | 45 (34.9%) | 106 (35.5%) | 151 (35.3%) |
| *Missing* | *2* | *1* | *0* | *3* | *3* |
| **EQ-5D anxiety & depression** | | | | | |
| No anxiety or depression present | 222 (71.2%) | 75 (64.1%) | 85 (65.9%) | 212 (70.4%) | 297 (69.1%) |
| Anxiety or depression present | 90 (28.8%) | 42 (35.9%) | 44 (34.1%) | 89 (29.6%) | 133 (30.9%) |
| *Missing* | *1* | *0* | *0* | *1* | *1* |
| **EQ-5D-5L index score** | | | | | |
| Median (IQR) | 0.89 (0.84 to 1.00) | 0.89 (0.85 to 1.00) | 0.89 (0.83 to 1.00) | 0.89 (0.85 to 1.00) | 0.89 (0.85 to 1.00) |
| Range | 0.07 to 1.00 | 0.41 to 1.00 | 0.44 to 1.00 | 0.07 to 1.00 | 0.07 to 1.00 |
| *Missing* | *3* | *1* | *0* | *4* | *4* |
| **EQ VAS** | | | | | |
| Median (IQR) | 85 (77 to 90) | 85 (77 to 90) | 85 (77 to 90) | 85 (77 to 90) | 85 (77 to 90) |
| Range | 20 to 100 | 40 to 100 | 50 to 100 | 20 to 100 | 20 to 100 |
| *Missing* | *8* | *2* | *5* | *5* | *10* |

*Legend: FAS = Fatigue Assessment Scale, mMRC = modified Medical Research Council, DASS-21 = Depression, Anxiety and Stress Score (21 items), EQ = EuroQol, VAS = Visual Analogue Scale, IQR = Interquartile Range.*

**Table S8**: Comparison of population characteristics of participants of the Zurich SARS-CoV-2 Cohort study and individuals not participating in the study.

| Variable | Participants included in analysis [a,b], N=431 | Enrolled Participants [b,c], N=442 | Nonparticipants [c], N=858 |
|---|---|---|---|
| **Age group (years)** | | | |
| 18-39 | 164 (38.1%) | 170 (38.4%) | 325 (37.9%) |
| 40-64 | 205 (47.6%) | 208 (47.1%) | 352 (41.0%) |
| ≥65 | 62 (14.4%) | 64 (14.5%) | 181 (21.1%) |
| **Sex** | | | |
| Female | 214 (49.7%) | 218 (49.3%) | 415 (48.4%) |
| Male | 217 (50.3%) | 224 (50.7%) | 440 (51.3%) |
| *Missing* | *0* | *0* | *3 (0.3%)* |
| **Initial symptoms** | | | |
| Symptomatic | 385 (89.3%) | 370 (83.7%) | 692 (80.7%) |
| Asymptomatic | 46 (10.7%) | 8 (1.8%) | 36 (4.2%) |
| *Missing* | *0* | *64 (14.5%)* | *130 (15.1%)* |
| **Time from symptom onset to diagnosis** | | | |
| Median (IQR) | - | 2 (1 to 5) | 2 (1 to 5) |
| **Hospitalization** | 81 (18.8%) | 58 (13.2%) | 205 (23.9%) |

*Legend: [a] Data presented refers to that collected in the Zurich SARS-CoV-2 Cohort study. [b] Data presented refers to that recorded in contact tracing files within the Department of Health. [c] Includes all participants who consented to participate in the Zurich SARS-CoV-2 Cohort study (i.e., including 6 individuals with suspected reinfection and 5 individuals that never filled a questionnaire). Time from symptom onset to diagnosis was not retrospectively elicited in our study.*
